# Forecasting laboratory measurements from longitudinal electronic health records

**DOI:** 10.64898/2026.08.12.26360243

**Authors:** Farzaneh Firoozbakht, Jan Baumbach

**Affiliations:** Institute for Computational Systems Biology, University of Hamburg, Albert-Einstein-Ring 8-10, 22761 Hamburg, Germany; Department of Mathematics and Computer Science, University of Southern Denmark, Odense 5000, Denmark.

## Abstract

Forecasting a patient’s laboratory measurements at future clinical visits from longitudinal electronic health records (EHRs) can support disease monitoring and treatment planning in the context of personalized medicine. However, accurate prediction remains challenging since patients exhibit complex and highly individualized clinical trajectories. Here, we present LaBERT, a transformer-based model trained to forecast future laboratory measurements of a patient given information available at the current and previous clinical visits. Evaluated on 583,535 clinical visits from 255,769 patients in the MIMIC-IV database, LaBERT consistently outperformed baseline methods, reducing mean squared error from 0.77 to 0.53 and improving the coefficient of determination (R^2^) from 0.29 to 0.51. Medication perturbation analysis further showed that LaBERT learns treatment-related information that is clinically meaningful. In particular, we showed that using the originally prescribed medications, LaBERT predicted future patient states more accurately than when using randomized medication sets in 81% of visits. Furthermore, our controlled counterfactual analyses reproduced established pharmacological effects, including warfarin-associated increases in international normalized ratio (INR) and heparin-associated increases in activated partial thromboplastin time (aPTT), consistently across multiple prediction horizons. These findings establish LaBERT as a model for forecasting future laboratory measurements from longitudinal EHRs and provide a foundation for treatment-dependent patient-state simulation and personalized clinical decision support.

## Introduction

Laboratory measurements are central to clinical decision-making, providing quantitative indicators of physiological state used to monitor disease progression, assess treatment response, and guide therapeutic decisions (Rajkomar et al. 2018). Forecasting how these measurements will change could provide early insight into a patient’s evolving physiological state. However, predicting future laboratory measurements is challenging due to the complex interactions among disease progression, therapeutic interventions, and individual patient history. Addressing this challenge requires large-scale longitudinal clinical data that capture these dynamics over time.

Advances in healthcare digitization have led to the widespread adoption of electronic health records (EHRs) across healthcare systems (Adler-Milstein et al. 2017), providing an increasingly rich source of longitudinal clinical data. By 2021, EHR adoption had reached 78% among physicians and 96% among acute care hospitals in the United States, representing a substantial increase over the previous decade (ASTP Health IT Research & Analysis 2022). The growing availability of large-scale clinical datasets, including the Medical Information Mart for Intensive Care (MIMIC), the eICU Collaborative Research Database (Pollard et al. 2018), HiRID (Faltys et al. 2021), UK Biobank (Allen et al. 2014), and the All of Us Research Program (All of Us Research Program Investigators et al. 2019), has created unprecedented opportunities to develop machine learning (ML) models that leverage longitudinal patient records for clinical prediction and decision support.

Recent advances in deep learning (DL) have enabled the analysis of longitudinal EHR data across a range of clinical prediction tasks, including mortality and clinical risk prediction (Hyland et al. 2020; Rong et al. 2025; Cummings et al. 2021), medication prediction (Alghamdi and Mostafa 2025), and the prediction of future clinical event and patient trajectory (Renc et al. 2024; Zhang et al. 2026). The latter approaches aim to predict or simulate subsequent events in a patient’s clinical course, including disease onset and progression, treatment response, and adverse events. Yet, comparatively little attention has been devoted to forecasting continuous laboratory measurements at future time points, where the objective is to provide a quantitative forecast of the patient’s physiological state. Recent work by Im et al. (Im et al. 2025) introduced LabTOP, an autoregressive transformer for laboratory value prediction trained on longitudinal EHRs. However, LabTOP focuses on short-term prediction from high-density ICU records, with limited applicability to sparse and irregular longitudinal data. To the best of our knowledge, no previous study has directly addressed forecasting continuous laboratory measurements at subsequent clinical visits.

In this study, we introduce LaBERT (Figure 1), a transformer-based framework trained on longitudinal EHRs to forecast continuous laboratory measurements at a queried future time point. Given a patient’s clinical history available up to a specific time point, including laboratory measurements, diagnoses, medications, procedures, and vital signs, together with the time interval to the future visit, LaBERT predicts 46 continuous clinical measurements (Supplementary Table 1).

**Fig. 1.**
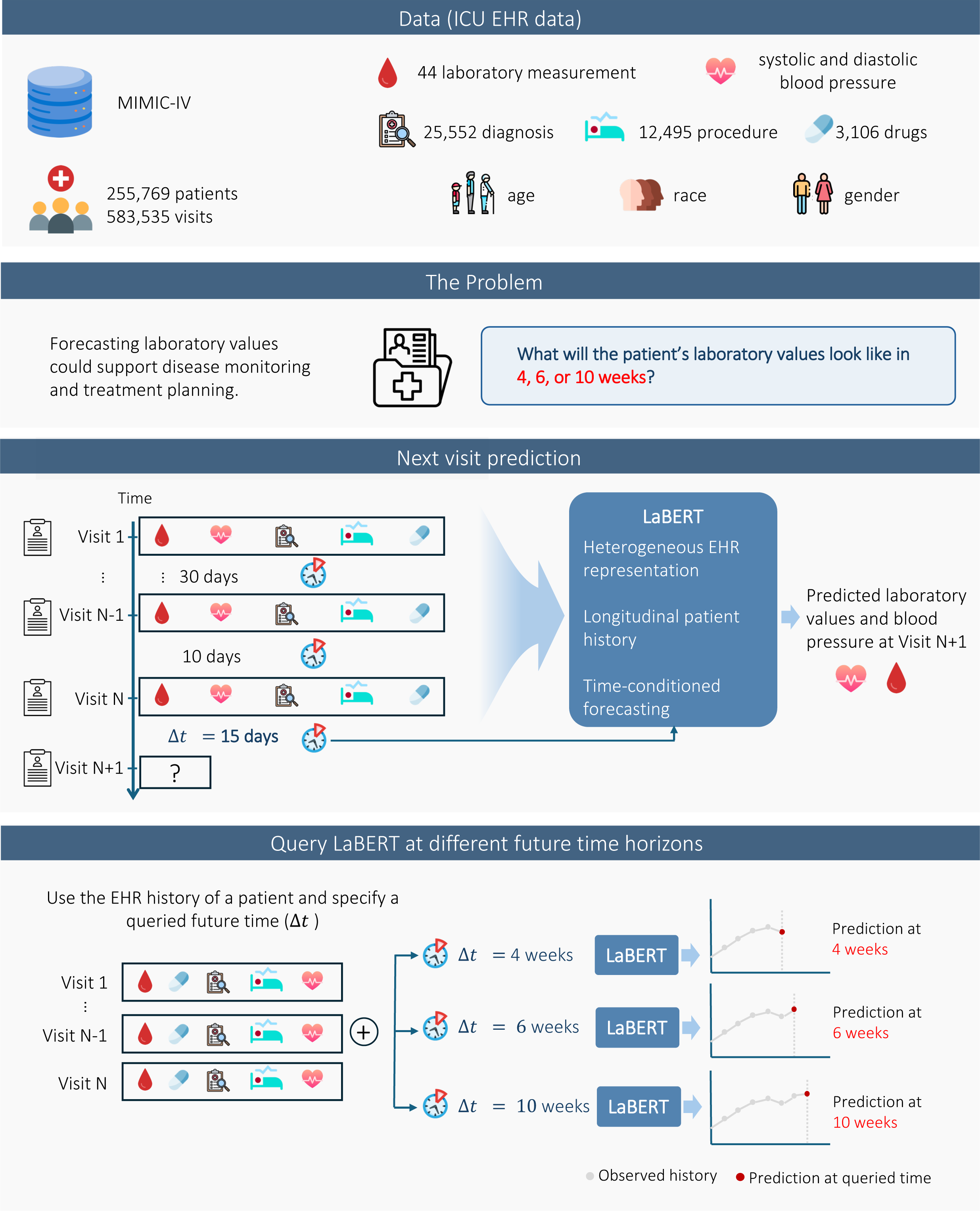
| Overview of LaBERT for forecasting future laboratory measurements from longitudinal EHRs. Raw data is obtained from MIMIC-IV that includes demographics, diagnoses, medications, procedures, and laboratory measurements. LaBERT uses heterogeneous clinical information from current and previous patient visits, including laboratory measurements, diagnoses, medications, procedures, and vital signs, together with the time interval to the subsequent visit (Δt), to predict continuous laboratory measurements and blood pressure at the next visit. During inference, the model can be queried with different future time intervals to forecast laboratory measurements at different future time points.

The main methodological contributions of this work are as follows:

- **Heterogeneous clinical representation.** LaBERT represents categorical clinical variables using learnable lookup embeddings and continuous physiological measurements using dedicated linear projections, preserving their numerical structure rather than discretizing or serializing all variables as textual tokens.
- **Feature-aware token representation.** Unlike conventional transformers, in which positional embeddings encode token order, LaBERT uses learnable feature-specific embeddings to encode the identity of each clinical variable. This enables heterogeneous laboratory measurements, diagnoses, medications, procedures, vital signs, and temporal variables to be jointly modeled, independent to their order, within a unified self-attention architecture.
- **Longitudinal patient modeling.** LaBERT propagates patient history as a latent representation, across successive clinical visits using a gated recurrent neural network architecture, enabling the model to capture temporal dependencies beyond individual patient encounters.
- **Time-conditioned forecasting.** By explicitly incorporating the interval between visits (t), LaBERT forecasts laboratory measurements at different future time horizons rather than being restricted to a fixed prediction interval.

We evaluated LaBERT using real-world clinical data from MIMIC-IV and demonstrated that it consistently outperforms baseline methods across multiple evaluation metrics. We further show that incorporating longitudinal patient history significantly improves predictive performance, highlighting the importance of modeling temporal dependencies across clinical visits. Furthermore, counterfactual analyses demonstrate that LaBERT reproduces the expected drug effects, including warfarin-associated increases in INR and heparin-associated increases in PTT, across multiple prediction horizons. These findings establish LaBERT as a framework for longitudinal patient-state forecasting and treatment-dependent trajectory simulation. To the best of our knowledge, no existing method combines heterogeneous clinical representations, feature-aware token modeling, explicit longitudinal history integration, and time-conditioned forecasting within a unified transformer framework for predicting future physiological state of patients.

## Method

### Dataset

In this study, we used the MIMIC-IV database (A. E. W. Johnson et al. 2023), a large publicly available EHR resource. MIMIC-IV contains de-identified, longitudinal clinical data from 299,712 patients across 431,231 hospital admissions at Beth Israel Deaconess Medical Center between 2008 and 2019. Despite its name, MIMIC-IV covers hospital-wide encounters rather than only intensive care admissions. We used records from the hospital module, including admissions, diagnoses, procedures, medications, laboratory results, and vital signs, covering patients from both general ward and ICU stays. These records capture both acute conditions leading to hospital admission and underlying chronic comorbidities. For each patient visit, we extracted the following data types (Fig. 1):

1. *Static patient information*: demographic variables, including gender, age, and race.
2. *Admissions*: information on hospital admission and discharge times for each visit.
3. *Diagnoses:* information of patients’ primary diagnoses and comorbid conditions encoded using ICD-9-CM and ICD-10-CM codes. In total, 25,552 unique diagnosis codes were included.
4. *Clinical procedures*: procedures performed during hospital visits encoded using ICD-9-PCS and ICD-10-PCS codes. In total, 12,495 unique procedure codes were included.
5. *Medication administrations*: treatments that patients received during hospital stays. Although a wide range of administration statuses is recorded, we restricted the analysis to medications that were actively administered to patients (e.g., administered, started or restarted), excluding prescriptions and records indicating non-administration. In total, 3,106 unique medications were included and represented using their generic drug names.
6. *Laboratory measurements*: laboratory test results for each patient. In our analysis, owing to substantial missing values and the lack of a consistent categorization scheme, discrete laboratory variables were excluded and the analysis was restricted to continuous lab measurements. Among these, we retained the 44 most frequently measured laboratory tests, each reported in at least 20% of patient-visit pairs, to ensure sufficient coverage and reliability (Supplementary Table 1).
7. *Vital signs*: systolic and diastolic blood pressure as indicators of patient physiological status.

### Data preprocessing

To standardize heterogeneous EHR data into structured longitudinal sequences, we converted the data into the Medical Event Data Standard (MEDS) format. MEDS represents each patient’s longitudinal record as a chronologically ordered sequence of clinical events, with each event defined by a patient identifier, timestamp, and clinical code, together with an associated value when applicable (e.g., the numerical value of a laboratory measurement).

Each visit was defined as the set of all clinical events (diagnoses, medications, procedures, laboratory measurements, and vital signs) sharing a hospital admission identifier, with its start and end times taken as the earliest and latest event timestamps within that visit, respectively. For each patient-visit pair, we then calculated the time interval (t) from the end of that visit to the start of the subsequent visit. We further excluded patient visits with t > 500 days, as such intervals would violate the assumption of temporal continuity in patient trajectories. Next, we retained laboratory measurements that are reported in at least 20% of patient-visit pairs (Supplementary Fig. 1). Following these preprocessing steps, the dataset comprised 583,535 visits for 255,769 patients as well as 50 features, including diagnoses, medications, procedures, time intervals between visits, systolic and diastolic blood pressure, and 44 laboratory measurements. On average, each patient has 2.3 hospital visits, with a minimum of 1 and a maximum of 125. The distribution of visit counts per patient is shown in Supplementary Fig. 1. Notably, 140,459 patients have only a single recorded visit, accounting for 55% of the cohort (Fig. 2a).

**Fig. 2.**
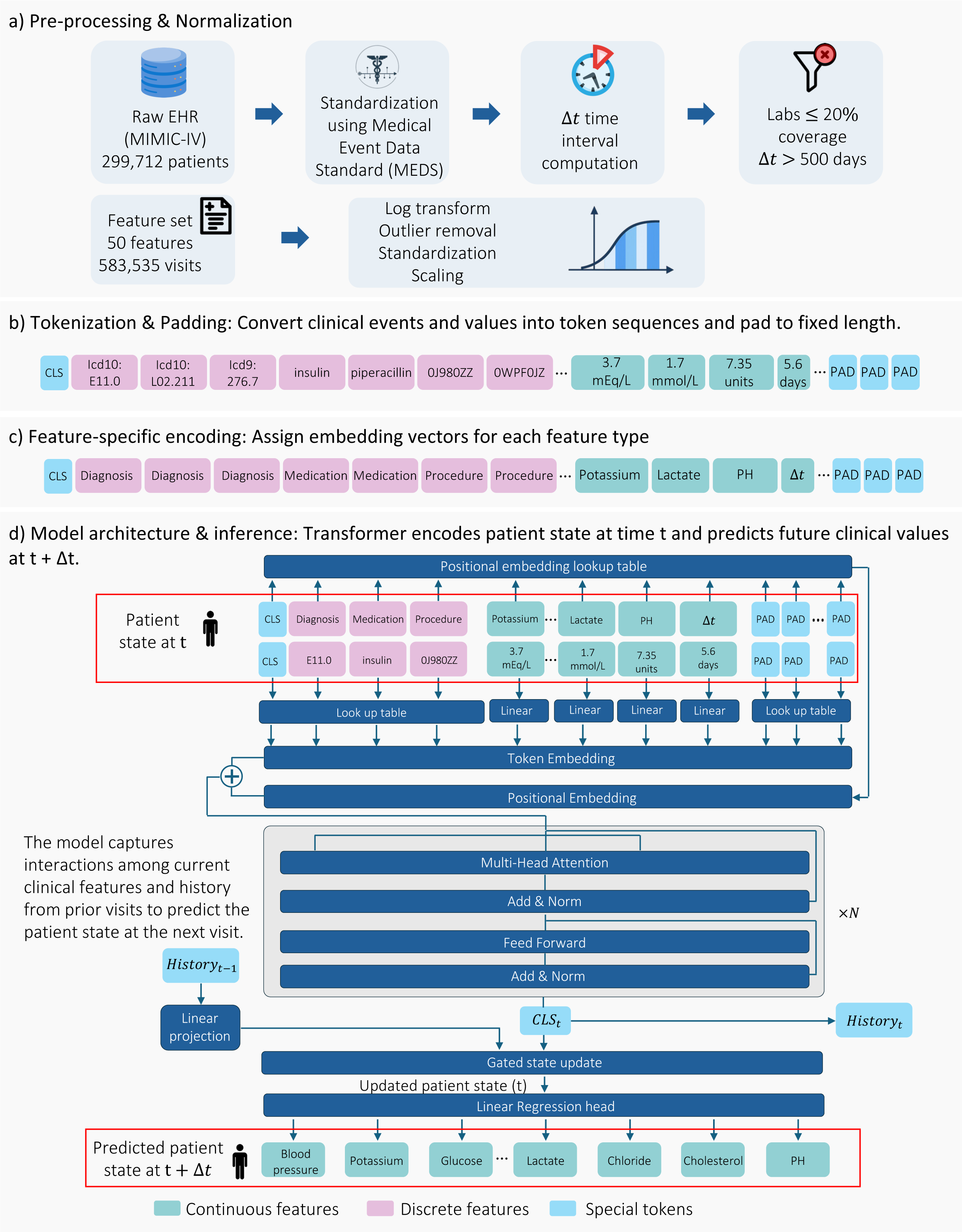
| Overview of LaBERT, our proposed pipeline for modeling longitudinal patient trajectories from ICU EHR data. (a) Pre-processing and normalization: events are standardized using MEDS, temporally aligned into fixed intervals, filtered for coverage, and numerically transformed (log scaling, outlier removal, normalization). (b) Tokenization and padding: heterogeneous clinical features are converted into sequences of discrete and continuous tokens with special tokens of CLS and PAD. (c) Positional encoding: tokens are defined based on clinical feature types. (d) Model architecture: Input embeddings are processed by a Transformer encoder to learn interactions across features; the CLS token summarizes the patient state at time t, which is used by a regression head to predict future clinical variables at time t + Δt.

### Data normalization

To normalize patient measurements, we computed the skewness of each feature and applied a log(1 + x) transformation to highly skewed variables (skewness > 2) after clipping negative values to zero. Features with skewness ≤ 2 were left on their original scale. Extreme outliers were then identified as those exceeding 10 median absolute deviations (MAD) from the median and considered as missing values. Finally, all variables were standardized to zero mean and unit variance. For time-related variables, age was scaled to the [0, 1] range using min-max normalization. t was log-transformed using a log (1 + x) function to reduce skewness and subsequently scaled to the [0, 1] range using min-max normalization (Fig. 2a).

### Sequence construction and feature-specific encoding

To prepare model inputs, clinical events and measurements were converted into sequences of tokens representing each patient visit. Each sequence contains discrete clinical events (i.e., diagnoses, medications and procedures) and continuous laboratory measurements (i.e., laboratory values, vital signs and t). Sequences were constructed with a fixed context length of 250 tokens corresponding to the maximum number of tokens observed across visits, and shorter sequences were padded using a special token to equalize the sequence length across all patients (Fig. 2b). In addition, we define a feature-specific encoding in which each token is assigned a learnable embedding corresponding to its feature identity rather than its position within the sequence. In particular, each token type (e.g. diagnosis, medication, blood pressure) is assigned a learnable vector; notably, all tokens belonging to the same type share a single positional vector for that entire category.

### Model architecture

Predicting future laboratory measurements from heterogeneous clinical data requires models capable of capturing complex relationships across heterogeneous clinical variables. LaBERT models the laboratory measurements at the subsequent clinical visit as a function of the current patient state, longitudinal patient history, and the time interval between visits:

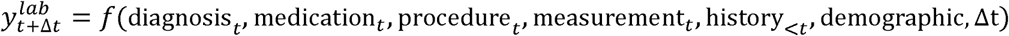

The model takes as input a sequence of clinical tokens representing a patient visit at time t and is trained to predict laboratory measurements *y* at time *t* +Δ*t*. Discrete features are represented using learnable embedding matrices, whereas each continuous feature is projected into the embedding space using a dedicated linear layer. In addition, each feature is assigned a learnable positional embedding, which is summed with its corresponding feature embedding (Fig. 2c). The resulting representations are processed by a multi-layer transformer encoder composed of stacked self-attention and feedforward layers with GELU activation functions (Vaswani et al. 2017). The contextual representation of the visit, encoded by the [*CLS*] token, is passed through a linear prediction head to estimate the patient’s laboratory state at the next clinical visit.

### Longitudinal history modeling

We extended LaBERT to incorporate longitudinal patient history when predicting laboratory measurements at the next clinical visit. To this end, patient visits were organized chronologically and processed sequentially by the model.

For visit, the transformer encoder produces contextualized embeddings, and the embedding corresponding to a designated summary token *CLS_n_* ∈ *R^d^* is extracted to form the visit-level representation. To model temporal dependencies across visits, we define a latent history state *h*_*n*-1_ ∈ *R^d^*, which summarizes information from all previous visits. At each step, the history state *h_n_* is updated by combining the current visit representation *CLS_n_* with the history state *h*_*n*-1_. Notably, the resulting state *h_n_* serves both as the representation used for prediction and as the history input for the next visit.

We investigated three methods for integrating history: (i) additive incorporation of a projected history vector into the current visit representation, (ii) a gated update mechanism that adaptively balances past and current information, and (iii) an LSTM-based update with explicit hidden and cell states. The three update mechanisms are defined as follows:

1. Additive integration, where the previous history *h_n_*_-1_ is linearly projected and added to the current visit representation :

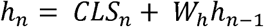 where *W_h_* ∈ *R^d^*^x*d*^.
2. Gated integration, where a learnable sigmoid gate (*Z_n_*) that adaptively balances the contributions from past and present representations:

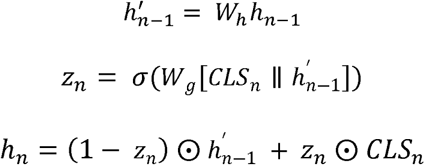 where || denotes vector concatenation and ⵙ element-wise multiplication.
3. LSTM-based integration, where the history is updated using input, forget, and output gates with an additional cell state:

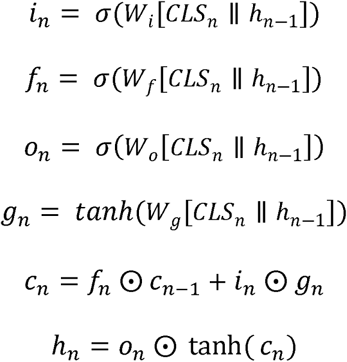

In this method, the history update introduces an explicit memory cell *c_n_* that allows the model to regulate information flow across visits. The input gate *i_n_* controls the contribution of the current visit representation *CLS_n_* to the memory *c_n_*, while the forget gate *f_n_* controls how much of the previous memory cell *c_n-_*_1_ is retained. The output gate *o_n_* further controls the flow of the updated memory to the hidden state *h_n_*. Together, these components enable the model to selectively retain, update, or discard historical information, allowing it to capture both short-term and long-term temporal dependencies in patient trajectories.

## Result

### Predictive performance

We evaluated model performance for the prediction of clinical measurements (i.e., laboratory and vital signs) for each patient-visit pair at their next-visit. After preprocessing, the dataset comprised 583,535 visits from 255,769 patients across 44 clinical features. Patients were splitted into train, validation, and test sets with ratios of 0.70, 0.15, and 0.15, respectively (Fig 3a). LaBERT was trained and evaluated across 11 runs with random seeds from 0 to 10. We benchmarked LaBERT against three baseline methods across test data:

[i] naive forecasting that predicts each measurement using its most recent prior value, reflecting the temporal persistence assumption commonly adopted in clinical time-series analysis. Results of the average performance on the test set show that LaBERT consistently outperforms the naive baseline across all evaluation metrics (Fig. 3). The model substantially reduces prediction error, decreasing MSE from 0.76 to 0.55 (Fig. 3b), corresponding to a reduction of approximately 28%, and root mean squared error (RMSE) from 0.87 to 0.74 (Supplementary Table 2). The coefficient of determination (R^2^) improves from 0.30 to 0.5, indicating that the model captures variation beyond simple temporal persistence (63% improvement). Improvements are also observed in correlation-based metrics, with global Pearson correlation increasing from 0.65 to 0.70 and average on per-laboratory correlation from 0.60 to 0.65 (Fig. 3c,d).
[ii] a multilayer perceptron (MLP) trained to predict laboratory values at the next visit from patient measurement. Our Comparison shows that LaBERT consistently outperforms the MLP across all evaluation metrics. In particular, LaBERT reduces prediction error, decreasing mean squared error from 0.60 to 0.55, corresponding to a reduction of approximately 8.3%, and root mean squared error from 0.77 to 0.74. R^2^ improves from 0.45 to 0.50 (11% improvement). Improvements are also observed in correlation-based metrics, with global Pearson correlation increasing from 0.67 to 0.70 and per-laboratory correlation from 0.60 to 0.65 (Supplementary Table 2). These improvements reflect the added contribution of token-level sequence modeling and attention mechanism.
[iii] LabTOP (Im et al., 2025), a GPT-2-based autoregressive model that represents clinical events as text tokens and predicts laboratory values as raw numeric text within an ICU stay. Although no directly comparable published methods currently exist for our prediction setting, we adapted LabTOP inference to serve as the closest available baseline. In its original formulation, LabTOP estimates a single laboratory value at each prediction step using all preceding medical events as context, with each ICU stay treated as an independent sample. In contrast, LaBERT simultaneously predicts multiple laboratory values at a patient’s next visit. To ensure a fair comparison, the LabTOP model was evaluated on a subset of test visits that were not observed during its training. Results show that LaBERT consistently outperforms LabTOP across all evaluation metrics, computed on the test set common to both models (N = 335,236 prediction pairs across 43 shared laboratory tests). LaBERT significantly improves R^2^ from −3.7 for LabTOP, indicating performance worse than predicting the population mean, to 0.77. Correlation-based metrics show a similar pattern, with global Pearson correlation increasing from 0.07 to 0.88 and mean per-laboratory Pearson correlation from approximately 0.00 to 0.60 (Supplementary Table 3).

**Fig. 3.**
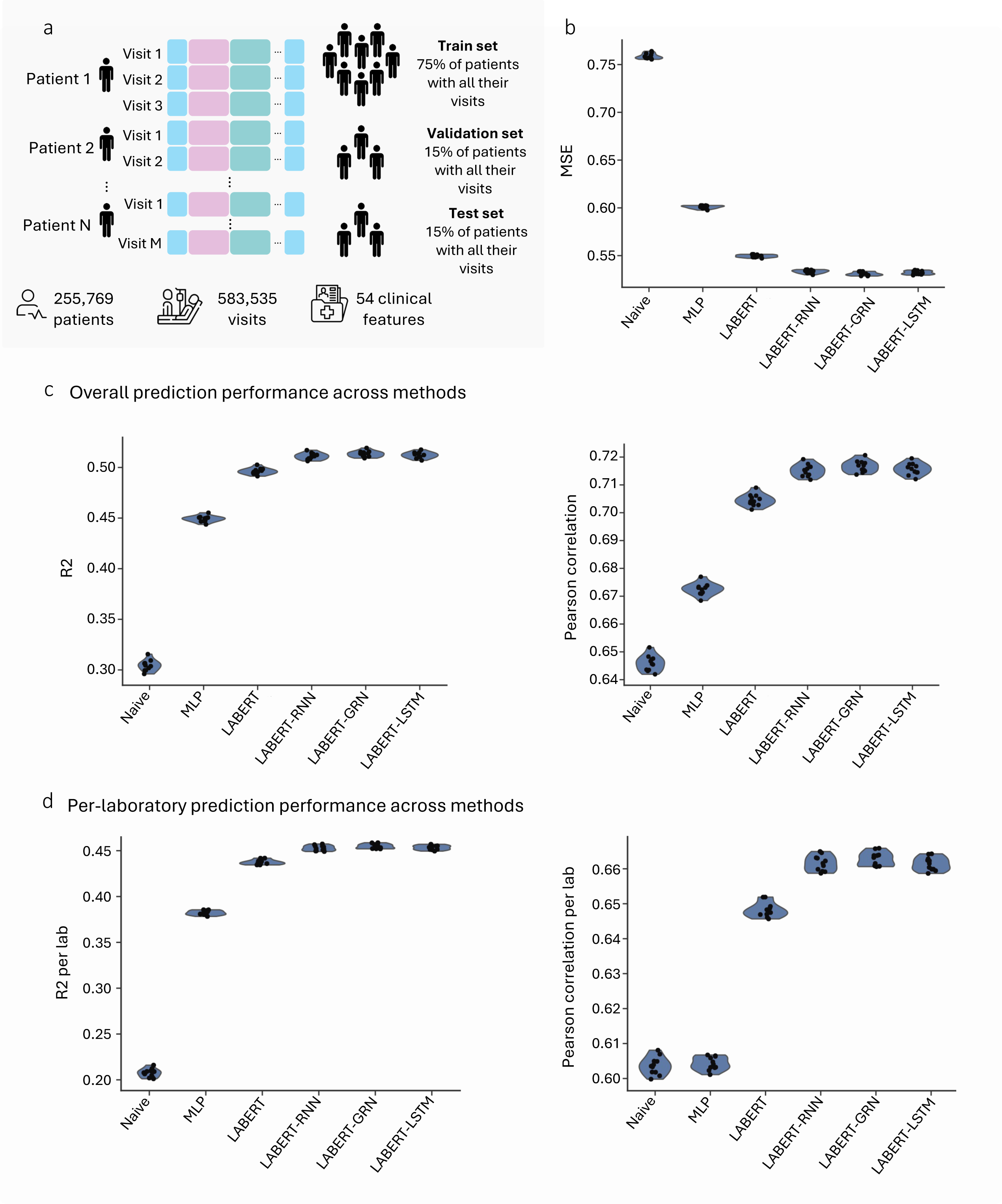
| Predictive performance of LaBERT. a) Patient trajectories are organized into longitudinal visits and split into training, validation, and test sets at the patient level. The dataset comprises 255,769 patients, 583,535 visits, and 54 clinical features. b) Prediction performance measured by mean squared error (MSE) across models. LaBERT achieves a 28% reduction in MSE compared to the baseline. c) Prediction performance in terms of R^2^ and Pearson correlation across methods. LaBERT substantially improves predictive accuracy, increasing R² by 69% and Pearson correlation by 10% relative to the baseline. d) Averaged per-laboratory prediction performance in terms of R^2^ and Pearson correlation. LaBERT demonstrates consistent gains across individual laboratory tests, with R^2^ improving by 147% and Pearson correlation by 8% compared to the baseline.

To assess whether additional model capacity improves predictive performance, we evaluated deeper embedding and prediction heads. Specifically, we replaced the single-layer continuous feature embedding with two- and three-layer feedforward networks (Supplementary Fig. 2) and likewise evaluated two- and three-layer decoder heads (Supplementary Fig. 3). Neither modification improved predictive performance (Supplementary Tables 4 and 5), indicating that the baseline architecture provides sufficient model capacity for this prediction task.

To evaluate the contribution of longitudinal history, we compared the base LaBERT model with three history-aware variants that incorporated information from previous visits using additive, gated, or LSTM-based integration. Incorporating visit history significantly improved predictive performance over the base model (Fig. 3 and Supplementary Table 2). Relative to the base model, all three variants significantly reduced prediction error (paired t-test, P < 0.0001), with MSE decreasing by 3.0% for additive integration, 3.2% for LSTM-based integration, and 3.4% for gated integration. Correspondingly, R^2^ increased by 3.0-3.4%, while global Pearson correlation improved by 1.5–1.7%. Notably, the gated integration achieved the best overall performance (MSE = 0.53, R^2^ = 0.51, global Pearson = 0.72), significantly outperforming both additive integration and LSTM-based integration (one-sided paired t-test; p-value <10^-4^). Although the performance differences between the three history-aware variants were modest, these results indicate that adaptive mechanisms for integrating historical information provide a small but consistent advantage over simple additive aggregation.

To evaluate the contribution of demographic variables, we performed an ablation analysis by incorporating age, sex, and race as additional input tokens. Including these variables did not improve predictive performance and instead resulted in a slight decrease across all evaluation metrics (Supplementary Fig. 4 and Supplementary Table 6). These findings indicate that explicitly incorporating demographic variables does not improve prediction beyond the longitudinal clinical features already captured by LaBERT. Consistent with this observation, our analysis showed that age and sex were associated with several laboratory measurements, with age correlated most strongly with urea nitrogen (ρ = 0.51) and RDW-SD (ρ = 0.41), and sex with creatinine (ρ = 0.38). In contrast, race showed only weak correlations across all clinical features (Supplementary Fig. 5). These results suggest that demographic information is either largely redundant with the longitudinal clinical trajectory or provides limited independent information for predicting clinical measurements at the next clinical visit. A detailed analysis of these relationships is provided in the Supplementary Information.

### Robustness against missing input laboratory information

Laboratory tests vary substantially in measurement frequency, with some tests performed routinely and others measured only sporadically, resulting in considerable differences in data availability across laboratory variables (Supplementary Fig. 1). Machine learning models often struggle to learn reliable patterns from limited data, suggesting that differences in laboratory test availability may influence prediction performance. We therefore investigated the relationship between laboratory test availability and prediction error to assess whether differences in measurement frequency bias model performance. We report the normalized root mean squared error (NRMSE), which scales prediction error by the variability of each laboratory measurement, allowing errors across different laboratory tests to be represented on a comparable scale.

As can be seen in Fig. 4a, no strong association between measurement frequency and NRMSE is observed. Only three routinely measured markers of mean corpuscular volume (MCV), mean corpuscular hemoglobin (MCH), and creatinine tend to exhibit lower prediction error. These variables exhibit strong inter-variable correlations and relatively stable temporal dynamics, likely contributing to their lower prediction error. Fig. 4b shows the pairwise correlation heatmap of the laboratory measurements. Accordingly, the strong correlations between MCV and MCH and between creatinine and Urea Nitrogen (0.83 and 0.68 respectively) facilitates the prediction of one measurement in the presence of the other. This observation is consistent with previous work by Luo et al. (Luo et al. 2016), who showed that correlations and redundancy among concurrently measured laboratory tests can support prediction of individual laboratory values. This behavior is also observed among less frequent measurements with low NRMSE. For instance, red cell distribution width (RDW) and red cell distribution width standard deviation (RDW-SD), which quantify related hematological properties and are strongly correlated, show low prediction performance despite RDW-SD has been measured in less than 40% of the cases.

**Fig. 4.**
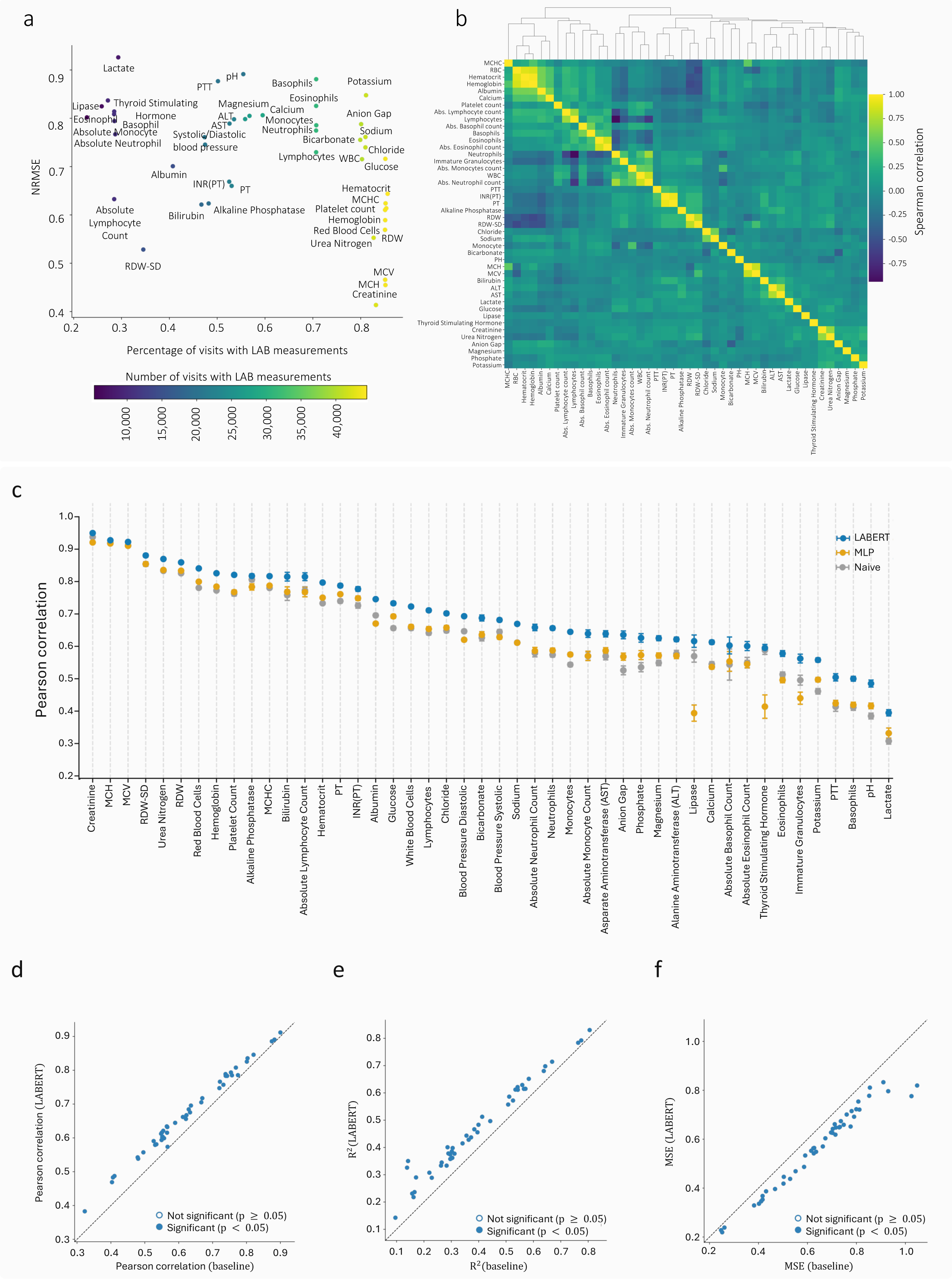
| Robustness to missing laboratory data and temporal modeling of patient trajectories. a) Normalized root mean squared error (NRMSE) across individual laboratory measurements against laboratory measurement availability. Each point represents one laboratory variable and is colored by their frequency in all visits. b) Pairwise Spearman correlation between laboratory measurements across the study cohort. Laboratory measurements are hierarchically clustered according to their correlation patterns. c) Pearson correlation between predicted and ground-truth values for each laboratory measurement, shown for LaBERT, the multilayer perceptron (MLP), and the naive baseline. Laboratory measurements are sorted by LaBERT performance. d-f) Comparison of LaBERT with the best-performing baseline (MLP or naive) across individual laboratory measurements using Pearson correlation between predicted and observed values, R^2^, and MSE, respectively. Each point represents one laboratory variable. Filled blue points indicate significant differences (paired t-test across seeds, P < 0.05). RBC: Red Blood Cells, ALT: Alanine Aminotransferase, WBC: White Blood Cells, and AST: Asparate Aminotransferase.

In contrast, physiologically dynamic biomarkers such as lactate, potassium, and coagulation parameters showed higher NRMSE. These variables are influenced by acute interventions and rapidly changing physiological states, making them less predictable from historical data. These results suggest that model performance remains challenging for rapidly evolving measurements.

Accordingly, our analysis suggests that lab measurement availability is not the primary determinant of predictive performance. Supporting this, closely related laboratory measurements with different availability can exhibit comparable prediction error. In addition, NRMSE for laboratory measurements with similar frequencies can cover a wide range from 0.4 to 1.0, supporting that lab measurement availability is not the primary determinant of predictive performance.

### Predictive performance across individual clinical measurements

In this section, we compare the predictive performance of the models across individual clinical measurements. To investigate this, we computed the Pearson correlation between predicted and observed values separately for each clinical measurement. As shown in Fig. 4c, LaBERT consistently outperformed the baseline methods across all individual clinical measurements. Pairwise comparisons using Pearson correlation, R^2^, and NRMSE showed that all clinical measurements exhibited significantly improved performance (P < 0.05). These findings indicate that the performance gains of LaBERT are broadly distributed across clinical measurements rather than being driven by a small subset of biomarkers. In addition, as discussed above, the high predictive accuracy observed for certain clinical measurements may be attributable to their relatively stable temporal dynamics. This is reflected in the strong performance of the naive forecasting baseline, which predicts each measurement using its most recent observed value. As temporal correlation decreases, the performance gap between LaBERT and the baseline models widens, indicating that LaBERT is particularly effective for clinical measurements with less predictable temporal dynamics.

### Patient trajectory forecasting across arbitrary time horizons

Predicting a patient’s laboratory measurement at different future time points could support clinical decision-making by enabling individualized monitoring and treatment planning. In MIMIC data, the interval between consecutive clinical visits varies across patients, ranging from 0 to 33,602 days (Supplementary Fig. 6). LaBERT explicitly incorporates the time interval between visits (Δ*t*) as an input, enabling prediction of patient state at arbitrary future time points. Supplementary Fig. 7 shows the average MSE across all clinical measurements as a function of Δ*t*. Prediction error remained relatively stable over a broad range of prediction horizons, from 1 day to 6 months, before increasing at longer intervals (> 6 months), indicating that LaBERT provides reliable forecasts across clinically relevant time scales.

To demonstrate the impact of Δ*t* as input, we compared the model’s performance to a baseline in which Δ*t* is fixed to zero for all patient visits (Fig. 5a). This setting corresponds to the naive temporal persistence assumption. Specifically, we define Δ*loss* as the difference in MSE between the standard model and the baseline (Fig. 5a). As shown in Fig. 5b, Δ*loss* is consistently positive (one-sided paired t-test, p-value < 10^-15^), indicating that setting Δ*t* to 0 increases the prediction error. Moreover, Δ*loss* increases with Δ*t*, demonstrating that the performance gap widens as the prediction horizon grows. For short time intervals, where clinical values tend to remain relatively stable, the contribution of Δ*t* is minimal, resulting in similar prediction errors with and without temporal information. In contrast, for longer intervals, omitting temporal information leads to substantially higher prediction error. These findings highlight the importance of Δ*t* for predictive performance and indicate that the model learns a temporally coherent representation of patient state, rather than relying solely on static patient features.

**Fig. 5.**
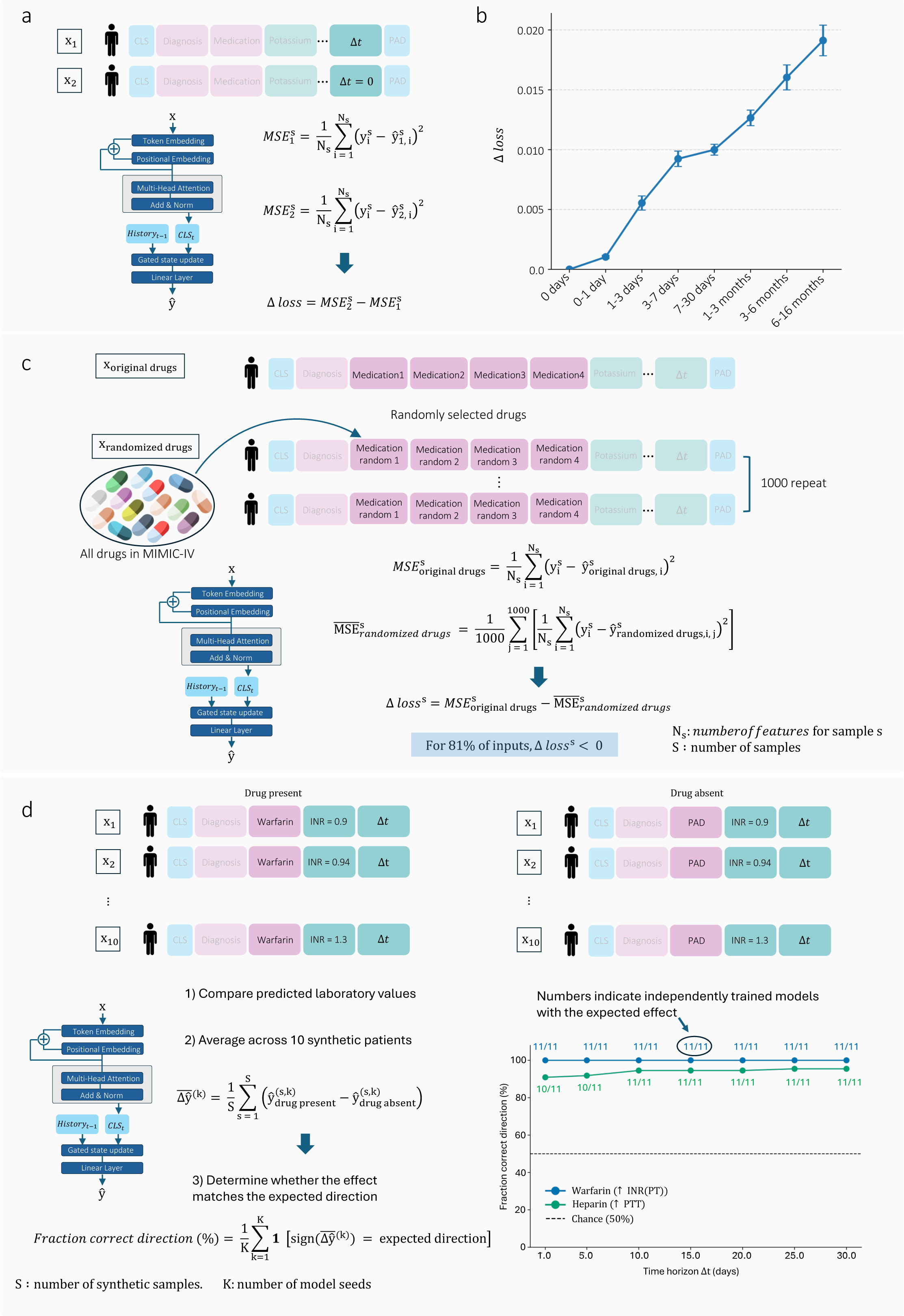
| Modeling temporal dynamics and treatment-dependent patient trajectories. a) Schematic of the temporal ablation analysis. Prediction performance of LaBERT using the time interval between visits (Δt) was compared with a model in which Δt was fixed to zero. Δloss denotes the difference in mean squared error (MSE) between the two settings. b) Δloss as a function of the interval between visits. Positive values indicate that replacing the true Δt with Δt = 0 increases prediction error. Error bars represent mean ± std across model seeds. c) Schematic of medication perturbation analysis. For each patient-visit pair, the original medications were replaced with randomly selected medications while preserving their number. The procedure was repeated 1,000 times, and prediction performance was compared with that obtained using the original medications. d) Schematic of counterfactual evaluation of drug-laboratory relationships. Synthetic patient profiles differing only in the presence or absence of a drug token were used to predict future clinical measurements. e) The fraction of independently trained LaBERT models reproducing the expected direction of the drug effect is shown across prediction horizons for warfarin-INR and heparin-PTT.

### Modeling treatment-dependent patient trajectories

A translational application of LaBERT is its ability to simulate patient trajectories under alternative treatment scenarios. The model predicts future clinical measurements following different medication interventions, enabling in silico evaluation of treatment-dependent changes in patient state. These predictions provide a framework for comparing alternative therapeutic strategies at the individual patient level.

To first assess whether LaBERT captures medication-specific information, we performed a medication perturbation analysis (Fig. 5c). The analysis was restricted to visits that (i) contained at least one recorded medication and (ii) were followed by a subsequent recorded visit, resulting in 18,495 visits (16% of the dataset). For each visit, we replaced the medication recorded at each visit with randomly selected medications, preserving the number of medications, and predicted the subsequent patient state. This procedure was repeated 1,000 times for each visit. As can be seen in Fig. 5c, the prediction loss obtained using the original medications was lower than the average loss across randomized medication sets in 81% of visits, indicating that LaBERT leverages medication information in predicting the future clinical states.

To further demonstrate the translational potential of LaBERT for clinical decision-making, we evaluated whether LaBERT reproduces established pharmacological effects in a controlled counterfactual setting. To this end, LaBERT was prompted to predict future clinical measurements for the same patient while varying only the medication input (Fig. 5d). This provides a controlled setting for evaluating whether LaBERT captures known drug effects: if the model has learned the pharmacological relationship, adding or removing the drug should shift the predicted clinical value in the expected direction. We specifically focused on drug-laboratory, where the drug effect is routinely monitored using the corresponding clinical measurement. Such relationships provide a more interpretable scenario for counterfactual evaluation, as changes in the monitored clinical measurement are directly linked to the drug’s known pharmacological action and are therefore less likely to be confounded by differences in underlying disease, patient characteristics, or other clinical factors. Accordingly, we identified the following two drug-clinical pairs:

1. Warfarin-INR: Warfarin inhibits vitamin K recycling, reducing the activation of vitamin K-dependent coagulation factors and thereby increasing INR.
2. Heparin-PTT: Heparin potentiates antithrombin-mediated inhibition of coagulation factors, particularly thrombin and factor Xa, thereby prolonging activated partial thromboplastin time (aPTT). Heparin potentiates antithrombin-mediated inhibition of coagulation factors, particularly thrombin and factor Xa, thereby prolonging activated partial thromboplastin time (aPTT).

For each pair, we constructed ten synthetic patient encounters that differed only in the baseline value of the target laboratory measurement. The ten values were evenly distributed across a predefined, clinically relevant range reflecting the context in which the corresponding drug is typically administered (Supplementary Methods). Each synthetic patient was presented to LaBERT to predict future clinical measurements at three time intervals (Δ *t*): 0.5, 2.5 and 30 days. For each encounter, we generated two otherwise identical input sequences that differed only in the drug input. In the drug-present condition, the sequence contained the corresponding drug token, whereas in the drug-absent condition, the drug token was replaced by a [*PAD*] token and therefore excluded by the model’s attention mask. Thus, the two conditions differed only in whether information about the drug was available to the model. We then compared the predicted value of the target laboratory measurement between the two conditions at each future time point to quantify the predicted drug-associated effect. To assess the robustness of these effects, we repeated each experiment across 11 independently trained GRN-based LaBERT models initialized with different random seeds.

The results, presented in Fig. 5d, were strongly consistent with the expected pharmacological effects. In the presence of warfarin, LaBERT predicted an increase in INR in 100% of comparisons across all 11 independently trained models and at every tested prediction horizon. Similarly, in the presence of heparin, LaBERT predicted the expected increase in PTT in 82-89% of comparisons, with consistent effects across 9-10 of the 11 models. Notably the predicted effects for both drug-laboratory pairs were consistent across forecast horizons ranging from 0.5 to 30 days, a 60-fold range, indicating that the observed relationships were robust to the choice of prediction horizon rather than being specific to a single evaluation setting.

## Discussion

Forecasting a patient’s future laboratory measurements from longitudinal EHR data could fundamentally improve the anticipation of disease progression, treatment response, and clinical decision-making. In this study, we introduced LaBERT, a transformer-based framework for predicting clinical measurements at subsequent clinical visits by jointly modeling diagnoses, medications, procedures, laboratory measurements, and the time interval between visits. Unlike previous approaches that primarily focus on predicting discrete clinical events or estimating patient risk, LaBERT directly forecasts continuous clinical measurements, providing a quantitative representation of future patient state. Across multiple evaluation metrics, LaBERT consistently outperformed baseline methods, demonstrating that our transformer-based architecture can capture more complex relationships underlying EHR data.

We showed that explicitly modeling longitudinal patient history improved predictive performance beyond representations derived from individual visits alone. All three history-aware variants consistently outperformed the history-free model, with gated integration achieving the strongest overall performance. These findings demonstrate that information from previous clinical encounters provides complementary information beyond the current visit, enabling more accurate forecasting of future patient states. The superior performance of gated integration further suggests that adaptively balancing historical and current information is more effective than treating all previous information equally.

We further showed that incorporating the time interval between visits enables LaBERT to forecast patient state across arbitrary prediction horizons. Removing Δ*t* consistently reduced prediction accuracy, with the impact becoming progressively larger as the interval between visits increased. These findings indicate that the model learns meaningful representations of temporal information that enable accurate forecasting across different prediction horizons. Notably, explicitly modeling Δ*t* enables LaBERT to generate predictions that are tailored to the expected time until the next clinical encounter.

Our analyses also provide insight into why certain laboratory measurements are easier to predict than others. Prediction accuracy was only weakly associated with measurement frequency, indicating that data availability alone does not explain model performance. Instead, laboratory measurements with high inter-variable correlations and relatively stable temporal dynamics were predicted more accurately, e.g. red blood cell indices and renal function markers. In contrast, rapidly changing measurements such as lactate and coagulation measurements, showed greater prediction error. These observations suggest that factors such as temporal stability of laboratory measurements and their relationships with other variables play a larger role than sampling frequency in determining prediction accuracy.

Beyond predictive accuracy, our results demonstrate that LaBERT learns clinically meaningful treatment-related information. Randomizing medication assignments consistently reduced prediction performance relative to the originally administered medications, indicating that the model captures treatment-dependent associations between medications and subsequent patient physiology. Furthermore, in controlled counterfactual experiments, LaBERT reproduced the known pharmacological effects of warfarin on INR and heparin on PTT across multiple prediction horizons. Because these experiments isolate medication identity while holding all other patient characteristics constant, they provide strong evidence that the model learns treatment-outcome relationships from longitudinal clinical data. Together, these findings establish a computational framework for simulating treatment-dependent patient trajectories and evaluating alternative therapeutic strategies in silico.

Although LaBERT captures longitudinal patient trajectories across a range of clinical measurements and prediction horizons, it is trained exclusively on observational EHR data and therefore learns associations present in routine clinical practice. Therefore, the presented results should not be interpreted as evidence that LaBERT estimates causal treatment effects. This phenomenon can be shown by insulin therapy. In the EHR data, patients receiving insulin exhibited significantly higher glucose levels at the subsequent visit than those not receiving insulin (200.7 mg/dL versus 192.5 mg/dL; P ≈ 7 × 10^-28^), despite having nearly identical baseline glucose levels (235.5 mg/dL versus 235.8 mg/dL; n = 8,950 and n = 17,171, respectively). Rather than contradicting the established glucose-lowering effect of insulin, this pattern likely reflects confounding by indication, as insulin is preferentially prescribed to patients with more severe or difficult-to-control hyperglycemia than is captured by a single baseline glucose measurement. Consistent with this observation, LaBERT reproduced the same pattern in counterfactual simulations, predicting higher future glucose values when insulin was included in otherwise identical synthetic patient profiles. These findings indicate that LaBERT faithfully captures treatment-outcome relationships present in observational EHR data, including clinically meaningful pharmacological signals as well as biases inherent in treatment assignment. Consequently, while LaBERT provides a useful framework for simulating treatment-dependent patient trajectories, its predictions should be interpreted as observational forecasts rather than causal estimates and should complement, rather than replace, clinical judgment or prospective evaluation. Future work could address this limitation by integrating longitudinal EHR representations with structured biomedical knowledge. Such biomedical knowledge could be represented using knowledge graphs, enabling longitudinal patient trajectories to be interpreted within the context of molecular mechanisms, disease biology, and drug actions (Firoozbakht et al. 2026). Combining observational patient trajectories with mechanistic biological knowledge may improve model interpretability, facilitate the identification of causal biological mechanisms underlying disease progression, and enable more biologically informed simulation of patient trajectories for personalized treatment planning.

## Code availability

All source code and documentation required to reproduce the results reported in this study will be made publicly available upon publication.

## Data availability

The MIMIC-IV database (v2.2) is publicly available through PhysioNet(A. Johnson et al. 2023) upon completion of the required human subjects research training and execution of a data use agreement. In accordance with the terms of this agreement, the raw patient-level data and derived datasets generated from MIMIC-IV cannot be redistributed.

## Author contributions

Conceptualization: F.F., J.B.

Methodology: F.F., J.B.

Investigation: F.F., J.B.

Visualization: F.F.

Software: F.F.

Funding acquisition: J.B.

Project administration: J.B.

Supervision: J.B.

Writing (Original draft): F.F.

Writing (Review & editing): F.F., J.B.

## References

Adler-Milstein, Julia, A. Jay Holmgren, Peter Kralovec, Chantal Worzala, Talisha Searcy, and Vaishali Patel. 2017. “Electronic Health Record Adoption in US Hospitals: The Emergence of a Digital ‘Advanced Use’ Divide.” Journal of the American Medical Informatics Association 24 (6): 1142–1148.

Alghamdi, Hanan, and Abeer Mostafa. 2025. “Advancing EHR Analysis: Predictive Medication Modeling Using LLMs.” Information Systems 131 (102528): 102528.

Allen, Naomi E., Cathie Sudlow, Tim Peakman, Rory Collins, and UK Biobank. 2014. “UK Biobank Data: Come and Get It.” Science Translational Medicine 6 (224): 224ed4.

All of Us Research Program Investigators, Joshua C. Denny, Joni L. Rutter, et al. 2019. “The ‘All of Us’ Research Program.” The New England Journal of Medicine 381 (7): 668–676.

ASTP Health IT Research & Analysis. 2022. “National Trends in Hospital and Physician Adoption of Electronic Health Records.” ASTP - Assistant Secretary for Technology Policy, March 24. https://healthit.gov/data/quickstats/national-trends-hospital-and-physician-adoption-electronic-health-records/?utm_source=chatgpt.com.

Cummings, Brandon C., Sardar Ansari, Jonathan R. Motyka, et al. 2021. “Predicting Intensive Care Transfers and Other Unforeseen Events: Analytic Model Validation Study and Comparison to Existing Methods.” JMIR Medical Informatics 9 (4): e25066.

Faltys, M., M. Zimmermann, X. Lyu, M. Hüser, and S. Hyland. 2021. “HiRID, a High Time-Resolution ICU Dataset (version 1.1. 1).” Physio. Net. https://scholar.google.com/citations?user=0FFcCdYAAAAJ&hl=en&authuser=1&oi=sra.

Firoozbakht, Farzaneh, Simon Suwer, Maria Louise Elkjaer, et al. 2026. “NetMedGPT - A Network Medicine Foundation Model for Extensive Disease Mechanism Mining and Drug Repurposing.” In bioRxiv. BioRxiv, January 4. 10.64898/2026.01.04.697552.

Hyland, Stephanie L., Martin Faltys, Matthias Hüser, et al. 2020. “Early Prediction of Circulatory Failure in the Intensive Care Unit Using Machine Learning.” Nature Medicine 26 (3): 364–373.

Im, Sujeong, Jungwoo Oh, and Edward Choi. 2025. “LabTOP: A Unified Model for Lab Test Outcome Prediction on Electronic Health Records.” In arXiv [cs.LG]. February 20. arXiv. 10.48550/arXiv.2502.14259.

Johnson, Alistair, Lucas Bulgarelli, Tom Pollard, Steven Horng, Leo Anthony Celi, and Roger Mark. 2023. “MIMIC-IV.” PhysioNet. 10.13026/6MM1-EK67.

Johnson, Alistair E. W., Lucas Bulgarelli, Lu Shen, et al. 2023. “MIMIC-IV, a Freely Accessible Electronic Health Record Dataset.” Scientific Data 10 (1): 1.

Luo, Yuan, Peter Szolovits, Anand S. Dighe, and Jason M. Baron. 2016. “Using Machine Learning to Predict Laboratory Test Results.” American Journal of Clinical Pathology 145 (6): 778–788.

Pollard, Tom J., Alistair E. W. Johnson, Jesse D. Raffa, Leo A. Celi, Roger G. Mark, and Omar Badawi. 2018. “The eICU Collaborative Research Database, a Freely Available Multi-Center Database for Critical Care Research.” Scientific Data 5 (1): 180178.

Rajkomar, Alvin, Eyal Oren, Kai Chen, et al. 2018. “Scalable and Accurate Deep Learning with Electronic Health Records.” Npj Digital Medicine 1 (1): 18.

Renc, Pawel, Yugang Jia, Anthony E. Samir, et al. 2024. “Zero Shot Health Trajectory Prediction Using Transformer.” Npj Digital Medicine 7 (1): 256.

Rong, Ruichen, Zifan Gu, Hongyin Lai, et al. 2025. “A Deep Learning Model for Clinical Outcome Prediction Using Longitudinal Inpatient Electronic Health Records.” In medRxiv. MedRxiv, January 23. 10.1101/2025.01.21.25320916.

Vaswani, Ashish, Noam Shazeer, Niki Parmar, et al. 2017. “Attention Is All You Need.” In arXiv [cs.CL]. June 12. arXiv. http://arxiv.org/abs/1706.03762.

Zhang, Andrew, Tong Ding, Sophia J. Wagner, et al. 2026. “A Multimodal and Temporal Foundation Model for Virtual Patient Representations at Healthcare System Scale.” In arXiv [cs.LG]. April 20. arXiv. 10.48550/arXiv.2604.18570.

