## Supplementary information for "Forecasting laboratory measurements from longitudinal electronic health records"

### Data

This study used the MIMIC-IV database (version 2.2) (Johnson et al. 2023), accessed through PhysioNet (Goldberger et al. 2000) under a data use agreement following completion of the required human subjects research training. Cohort selection and data preprocessing are described in the Method section and below.

### Hyperparameter optimization

To identify the optimal hyperparameters, we used Ray Tune (v2.49.2) (Liaw et al. 2018) to search a predefined hyperparameter space, with validation mean squared error (MSE) as the optimization objective. The search considered learning rate ($10e - 5, 10e - 4$), weight decay ( $10e - 6, 10e - 5, 10e - 4$), batch size (256, 512, 1024), hidden dimension (128, 256), number of attention heads (4, 8), and number of transformer encoder layers (2, 4, 6). A total of 40 hyperparameter configurations were evaluated using a patient-level training/validation/test split of 70/10/20. Each configuration was trained for up to 100 epochs using the Asynchronous Successive Halving Algorithm (ASHA) scheduler, which adaptively allocated computational resources and performed early stopping. The best-performing configuration consisted of a learning rate of $2.9 \times 10^{-5}$, weight decay of $1.7 \times 10^{-4}$, batch size of 512, hidden dimension of 256, eight attention heads, and six transformer encoder layers, and was used for all subsequent experiments.

### Implementation details

LABERT was implemented in Python (v3.12) using PyTorch (v2.7). The model consists of a transformer encoder with multi-head self-attention and GELU activation functions. Continuous clinical features were embedded using feature-specific linear projection layers, whereas diagnosis, procedure, and medication codes were represented using a shared learnable embedding matrix. The resulting token embeddings were processed by the transformer encoder, and the contextual representation of the [CLS] token was passed through a linear output layer to jointly predict all target laboratory measurements.

Model parameters were optimized using the AdamW optimizer with a cosine learning-rate schedule and linear warmup over the first 10% of training steps. The training objective was the mean squared error (MSE) computed only over observed laboratory measurements, with missing values excluded from the loss calculation. Training, validation, and test splits were constructed at the patient level to prevent information leakage between data partitions.

### Model architecture ablation

We evaluated whether increasing the complexity of the feature encoder improved predictive performance by comparing architectures with one to three feedforward transformation layers for each feature type (Table S4). Across all configurations, performance remained nearly identical, with only negligible differences across evaluation metrics. These findings indicate that a single linear projection is sufficient for embedding the input features and that predictive performance is primarily determined by the downstream transformer.

We performed an analogous analysis for the output decoder by varying the number of transformation layers used to map the transformer representation to the predicted laboratory values. Similarly, increasing decoder depth did not significantly affect performance, indicating that additional complexity in the output projection is unnecessary (Table S5).

### Baseline models

As a non-sequential baseline, we implemented a multilayer perceptron (MLP) using the same clinical information available to LABERT at each visit. Each visit was represented as a fixed-length feature vector comprising (i) 50 continuous variables (47 laboratory measurements, two vital signs, and the time interval to the subsequent visit (Δt)), with missing values imputed as zero, and (ii) a multi-hot binary vector encoding diagnosis, procedure, and medication codes from a shared vocabulary of 40,470 tokens. The resulting representation was processed by a feedforward neural network with GELU activation functions and a linear output layer that jointly predicted all 44 target laboratory measurements. Each visit was treated independently, without access to previous visits.

The model was trained using AdamW with a peak learning rate of $3 \times 10^{-4}$, linear warmup, cosine learning-rate decay, weight decay of $10^{-4}$, and a batch size of 5,000 for up to 200 epochs, with early stopping based on validation loss (patience = 5 epochs). Hyperparameter optimization was performed over batch size (5,000-40,000), hidden dimension (512 or 1,024), and network depth (two to five hidden layers). The best-performing configuration (hidden dimension = 512, three hidden layers, batch size = 5,000) was selected based on validation performance and subsequently evaluated across 11 random seeds using the same patient-level training, validation, and test splits as LABERT.

#### Adaptation of LabTOP

LabTOP was originally developed for a different prediction task than LABERT, generating a single laboratory value as free text from preceding clinical events within the same ICU stay. To enable comparison, we adapted only its inference procedure while leaving the published model architecture and training strategy unchanged.

Both models were evaluated on the same held-out patient cohort. Because LabTOP expects raw numerical values in textual form rather than normalized numerical inputs, the original unprocessed EHR records were used as its input. For each visit, LabTOP received all clinical events observed up to and including the current visit and was prompted to generate each laboratory value for the subsequent visit, thereby aligning its prediction target with that of LABERT while preserving its original autoregressive generation mechanism.

To prevent information leakage, patients included in the publicly released LabTOP training set were excluded from the evaluation cohort. Consequently, neither method was evaluated on patients observed during LabTOP training. Predictive performance was compared only on patient-visit laboratory pairs for which valid predictions were available from both methods.

### Analysis of demographic variables

To investigate why explicitly incorporating demographic variables did not improve predictive performance, we computed the Spearman correlation between each demographic variable and the continuous clinical features across the study cohort. Age and sex showed substantial, clinically expected correlations with a subset of laboratory measurements. Age correlated most strongly with urea nitrogen ($\rho = 0.51$) and RDW-SD ($\rho = 0.41$), consistent with age-related changes in renal function and hematological parameters, whereas sex correlated most strongly with creatinine ($\rho = 0.38$), reflecting known differences in muscle mass. In contrast, race showed uniformly weak correlations with all clinical variables. The strongest observed association (Black/African American with MCH) reached only $\rho = 0.17$, and the mean absolute correlation remained below 0.06 across all race categories.

### Case study selection for synthetic patient profiles

For both case studies, baseline laboratory values were selected to reflect the clinical context in which the corresponding drug is typically initiated. Warfarin is prescribed for conditions such as atrial fibrillation, and heparin for venous thromboembolism, where the monitored laboratory measurements, international normalized ratio (INR) and activated partial thromboplastin time (aPTT), respectively, are used to monitor the anticoagulant effect rather than to diagnose the underlying disease. Accordingly, synthetic baseline values for both drug-laboratory pairs were sampled from the normal reference range, with the expected drug effect represented as a shift away from baseline toward therapeutic anticoagulation.

### Hardware specifications

All experiments were conducted on a high-performance computing node equipped with four NVIDIA H200 NVL GPUs (each 143 GB).

*Table S1 | Laboratory measurements included in the study.*

MIMIC-IV item IDs and corresponding laboratory measurement names for the 44 laboratory variables retained after preprocessing.

| MIMIC-IV item ID | Laboratory measurement |
|---|---|
| 52069 | Absolute Basophil Count |
| 52073 | Absolute Eosinophil Count |
| 51133 | Absolute Lymphocyte Count |
| 52074 | Absolute Monocyte Count |
| 52075 | Absolute Neutrophil Count |
| 50861 | Alanine Aminotransferase (ALT) |
| 50862 | Albumin |
| 50863 | Alkaline Phosphatase |
| 50868 | Anion Gap |
| 50878 | Asparate Aminotransferase (AST) |
| 51146 | Basophils |
| 50882 | Bicarbonate |
| 50885 | Bilirubin, Total |
| 50893 | Calcium, Total |
| 50902 | Chloride |
| 50912 | Creatinine |
| 51200 | Eosinophils |
| 50931 | Glucose |
| 51221 | Hematocrit |
| 51222 | Hemoglobin |
| 51237 | INR(PT) |
| 52135 | Immature Granulocytes |
| 50813 | Lactate |
| 50956 | Lipase |
| 51244 | Lymphocytes |
| 51248 | MCH |
| 51249 | MCHC |
| 51250 | MCV |
| 50960 | Magnesium |

| | |
|---|---|
| 51254 | Monocytes |
| 51256 | Neutrophils |
| 51274 | PT |
| 51275 | PTT |
| 50970 | Phosphate |
| 51265 | Platelet Count |
| 50971 | Potassium |
| 51277 | RDW |
| 52172 | RDW-SD |
| 51279 | Red Blood Cells |
| 50983 | Sodium |
| 50993 | Thyroid Stimulating Hormone |
| 51006 | Urea Nitrogen |
| 51301 | White Blood Cells |
| 51491 | pH |

***Table S2 | Overall predictive performance of LABERT and baseline methods.***

Comparison of LABERT, the naive persistence baseline, and the multilayer perceptron (MLP) across the test set using mean squared error (MSE), root mean squared error (RMSE), coefficient of determination ($R^2$), global Pearson correlation, and mean per-laboratory Pearson correlation. Results are reported as the mean ± standard deviation across 11 independently trained models.

| | MSE | RMSE | MAE | R2 | R2 per LAB | Pearson Correlation | Pearson Correlation per LAB |
|---|---|---|---|---|---|---|---|
| **Baseline** | 0.75± 0.003 | 0.86± 0.002 | 0.60± 0.001 | 0.32± 0.005 | 0.24± 0.005 | 0.65± 0.003 | 0.62± 0.003 |
| **MLP** | 0.60± 0.002 | 0.77± 0.001 | 0.56± 0.001 | 0.45± 0.003 | 0.39± 0.003 | 0.68± 0.002 | 0.62± 0.002 |
| **LABERT** | 0.55± 0.001 | 0.74± 0.001 | 0.53± 0.001 | 0.50± 0.003 | 0.45± 0.003 | 0.71± 0.002 | 0.66± 0.002 |
| **LABERT-RNN** | 0.53± 0.002 | 0.73± 0.001 | 0.52± 0.001 | 0.52± 0.003 | 0.46± 0.003 | 0.72± 0.002 | 0.67± 0.002 |
| **LABERT-GRN** | 0.53± 0.002 | 0.73± 0.001 | 0.52± 0.001 | 0.52± 0.003 | 0.47± 0.003 | 0.72± 0.002 | 0.67± 0.002 |
| **LABERT-LSTM** | 0.53± 0.002 | 0.73± 0.001 | 0.52± 0.001 | 0.52± 0.003 | 0.47± 0.003 | 0.72± 0.002 | 0.67± 0.002 |

*Table S3 | Comparison of LABERT and LabTOP.*

Comparison of LABERT and LabTOP on the common evaluation set across 43 shared laboratory measurements. Performance is reported using MSE, RMSE, $R^2$, global Pearson correlation, and mean per-laboratory Pearson correlation.

| Measure | LabTOP | LABERT |
|---|---|---|
| $R^2$ | -3.7 | 0.77 |
| $R^2$ per LAB | -29750 | 0.38 |
| Pearson Correlation | 0.07 | 0.88 |
| Pearson Correlation per LAB | -0.004 | 0.60 |

*Table S4 | Ablation of continuous feature embedding architecture.*

Predictive performance obtained using one-, two-, and three-layer feedforward networks for embedding continuous clinical features. Results demonstrate that increasing embedding depth does not improve predictive performance.

| Measure | LABERT MLP-1L input | LABERT MLP-2L input | LABERT MLP-3L input |
|---|---|---|---|
| MSE | 0.550 ± 0.001 | 0.553 ± 0.003 | 0.557 ± 0.002 |
| RMSE | 0.741 ± 0.001 | 0.744 ± 0.002 | 0.746 ± 0.001 |
| MAE | 0.534 ± 0.001 | 0.535 ± 0.001 | 0.537 ± 0.001 |
| R2 | 0.496 ± 0.003 | 0.493 ± 0.004 | 0.490 ± 0.004 |
| R2 per LAB | 0.437 ± 0.003 | 0.434 ± 0.004 | 0.430 ± 0.004 |
| Pearson Correlation | 0.705 ± 0.002 | 0.703 ± 0.003 | 0.701 ± 0.002 |
| Pearson Correlation per LAB | 0.648 ± 0.002 | 0.647 ± 0.003 | 0.645 ± 0.002 |

*Table S5 | Ablation of prediction head architecture.*

Predictive performance obtained using one-, two-, and three-layer prediction heads. Results demonstrate that increasing decoder depth does not improve predictive performance compared with a single linear prediction head.

| Measure | LABERT 1L output | LABERT 2L output | LABERT 3L output |
|---|---|---|---|
| MSE | 0.550 ± 0.001 | 0.554 ± 0.002 | 0.555 ± 0.002 |
| RMSE | 0.741 ± 0.001 | 0.744 ± 0.001 | 0.745 ± 0.001 |
| MAE | 0.534 ± 0.001 | 0.536 ± 0.001 | 0.537 ± 0.001 |

| | | | |
|---|---|---|---|
| **R2** | 0.496 ± 0.003 | 0.492 ± 0.003 | 0.491 ± 0.003 |
| **R2 per LAB** | 0.437 ± 0.003 | 0.433 ± 0.003 | 0.431 ± 0.003 |
| **Pearson Correlation** | 0.705 ± 0.002 | 0.702 ± 0.002 | 0.701 ± 0.002 |
| **Pearson Correlation per LAB** | 0.648 ± 0.002 | 0.645 ± 0.002 | 0.644 ± 0.002 |

***Table S6 | Effect of demographic variables on predictive performance.***

Predictive performance of LABERT with and without demographic variables (age, sex, and race) included as additional input tokens. Performance is reported using MSE, RMSE, $R^2$, global Pearson correlation, and mean per-laboratory Pearson correlation, demonstrating that explicit inclusion of demographic variables does not improve predictive performance.

| **Measure** | **LABERT-GRN** | **LABERT-GRN with demographic features** |
|---|---|---|
| **MSE** | 0.531 ± 0.002 | 0.532 ± 0.002 |
| **RMSE** | 0.729 ± 0.001 | 0.729 ± 0.001 |
| **MAE** | 0.523 ± 0.001 | 0.520 ± 0.001 |
| **R2** | 0.513 ± 0.003 | 0.506 ± 0.003 |
| **R2 per LAB** | 0.455 ± 0.003 | 0.454 ± 0.002 |
| **Pearson Correlation** | 0.717 ± 0.002 | 0.712 ± 0.002 |
| **Pearson Correlation per LAB** | 0.663 ± 0.002 | 0.661 ± 0.002 |

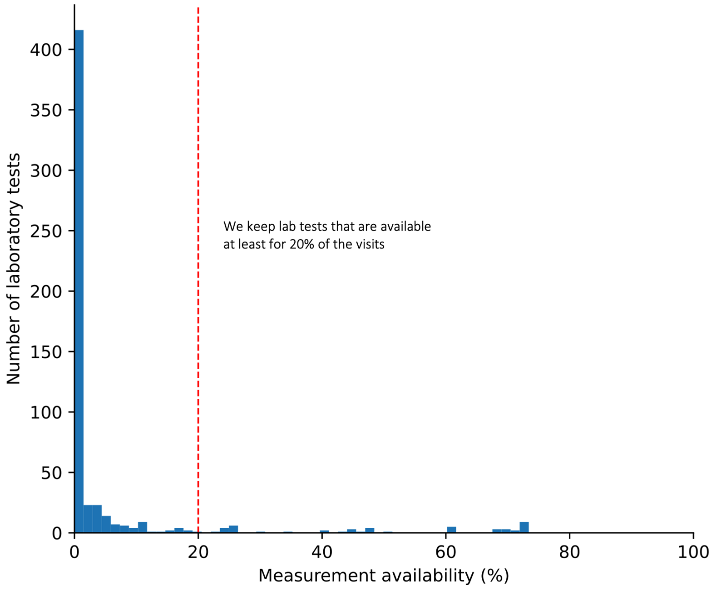

**Fig. S1 | Availability of laboratory measurements across patient visits.**

Distribution of laboratory measurement availability, defined as the percentage of patient visits in which each laboratory measurement was recorded. The red dashed line indicates the 20% availability threshold used for feature selection. Laboratory measurements recorded in fewer than 20% of visits were excluded, resulting in a final set of 44 laboratory variables.

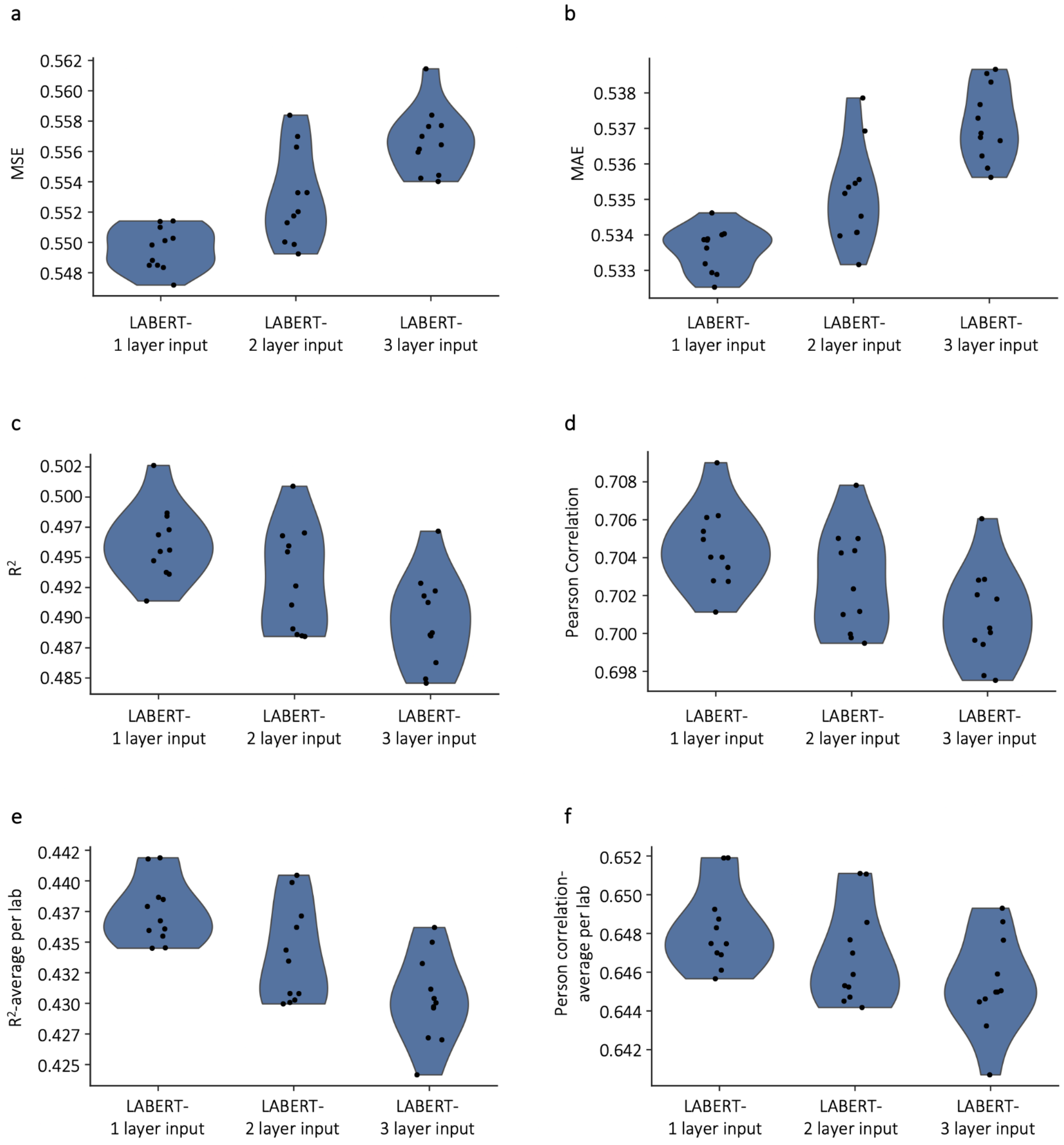

**Fig. S2 | Ablation of continuous feature embedding depth.**

Predictive performance of LABERT using one-, two-, or three-layer feedforward networks to embed continuous clinical features. (a) MSE. (b) MAE. (c) $R^2$. (d) Pearson correlation. (e) Mean per-laboratory $R^2$. (f) Mean per-laboratory Pearson correlation. Violin plots summarize results across 11 independently trained models; points indicate individual random seeds.

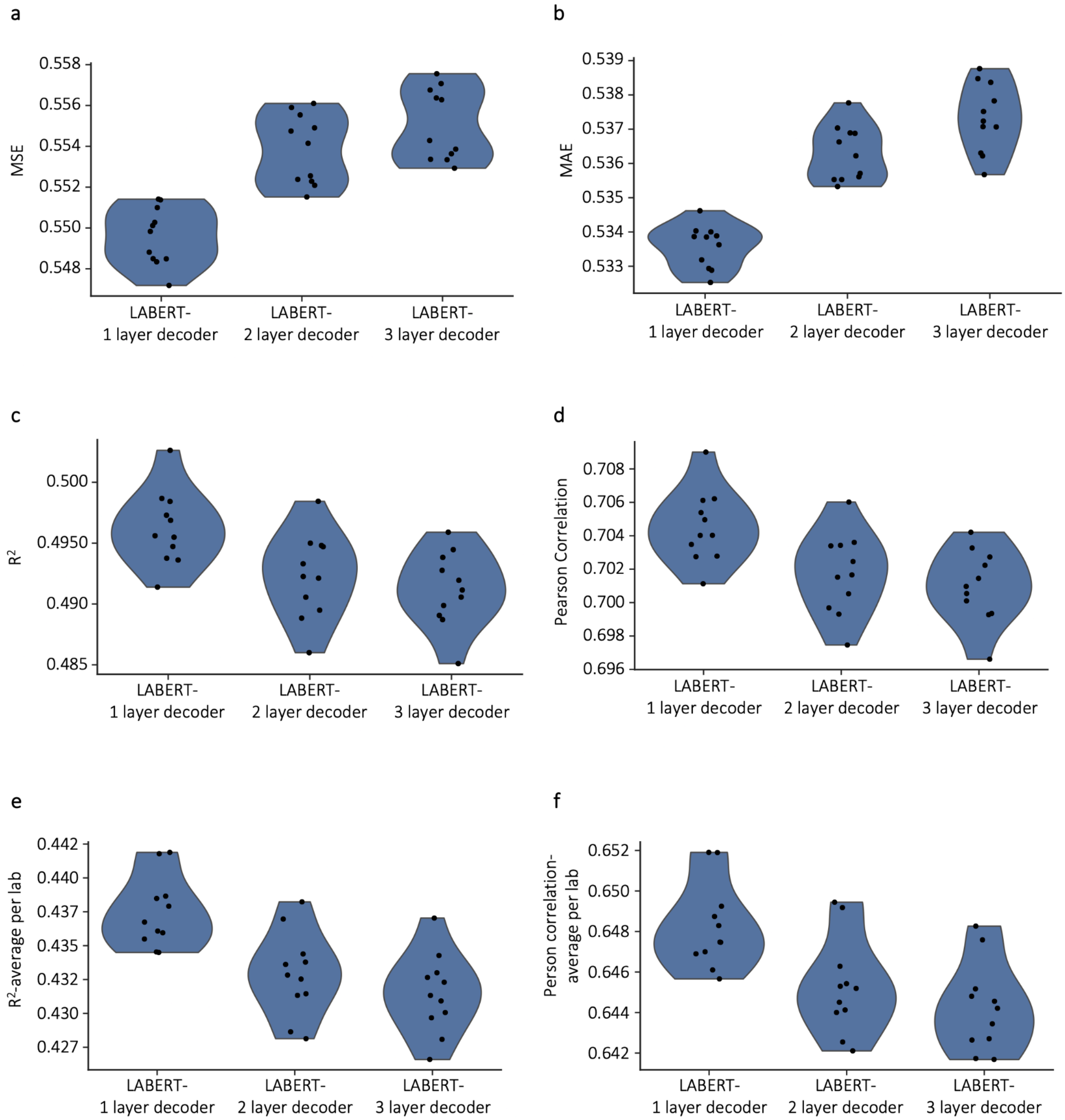

**Fig. S3 | Ablation of prediction head depth.**

Predictive performance of LABERT using one-, two-, or three-layer feedforward prediction heads. (a) MSE. (b) MAE. (c) $R^2$. (d) Pearson correlation. (e) Mean per-laboratory $R^2$. (f) Mean per-laboratory Pearson correlation. Violin plots summarize results across 11 independently trained models; points indicate individual random seeds.

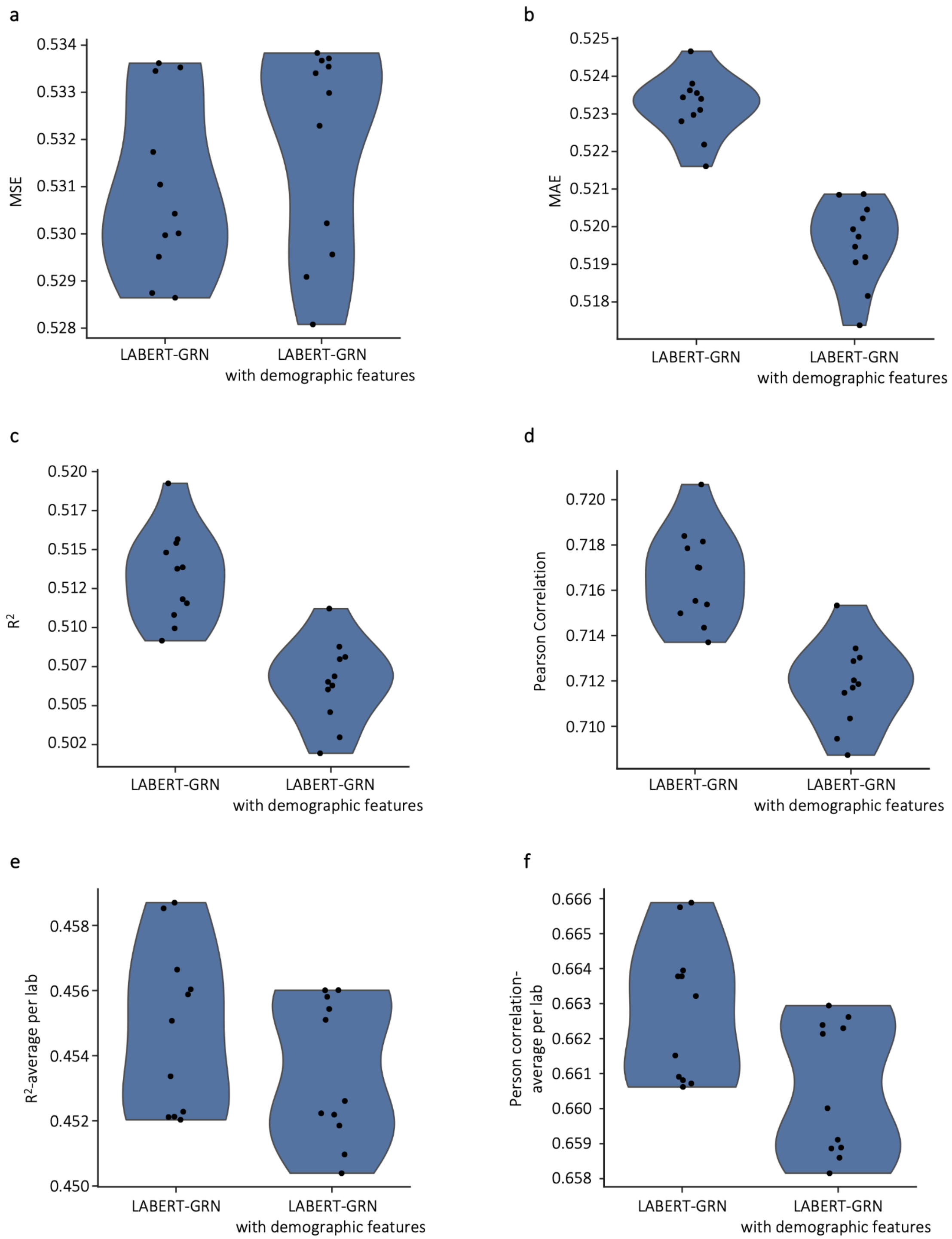

**Fig. S4 | Effect of demographic variables on LABERT performance.**

Predictive performance of LABERT-GRN with and without demographic variables (age, sex, and race) included as additional input tokens. (a) MSE. (b) MAE. (c) $R^2$. (d) Pearson correlation. (e) Mean per-laboratory $R^2$. (f) Mean per-laboratory Pearson correlation. Violin plots summarize results across 11 independently trained models; points indicate individual random seeds.

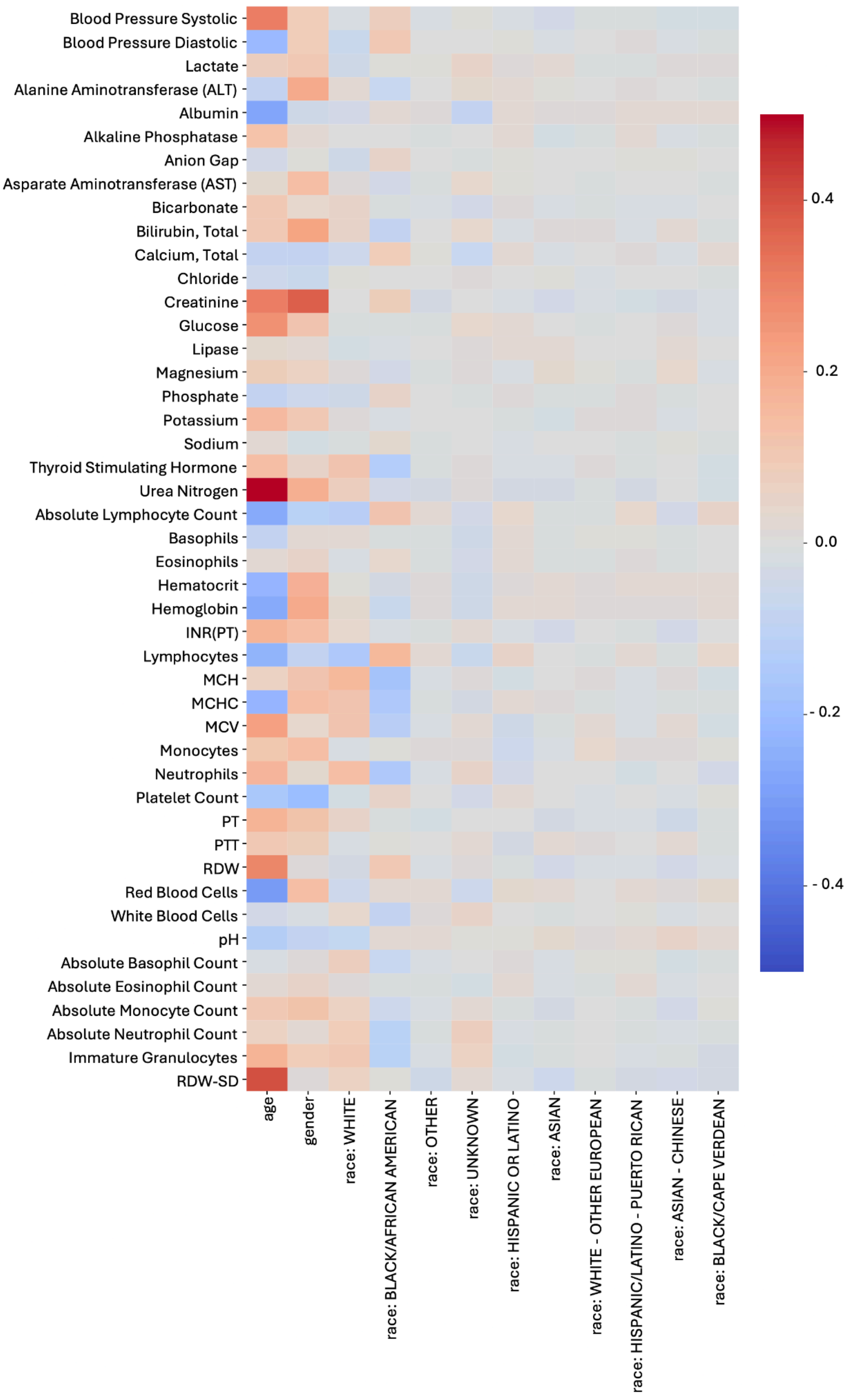

**Fig. S5 | Correlation between demographic variables and clinical features.**

Spearman correlation coefficients between demographic variables (age, sex, and the ten most frequent race categories) and clinical features across the study cohort. Age and sex show moderate correlations with a subset of laboratory measurements, whereas race exhibits uniformly weak correlations across clinical features.

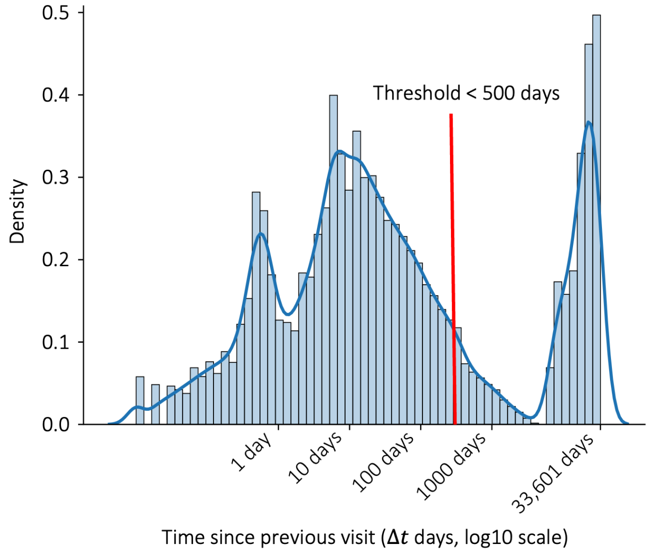

**Fig. S6 | Distribution of time intervals between consecutive clinical visits.**

Distribution of the time interval ($\Delta t$) between consecutive visits for the same patient, shown on a $log_{10}$ scale. The red dashed line indicates the exclusion threshold of $\Delta t$ = 500 days; visit pairs with longer intervals were excluded from subsequent analyses.

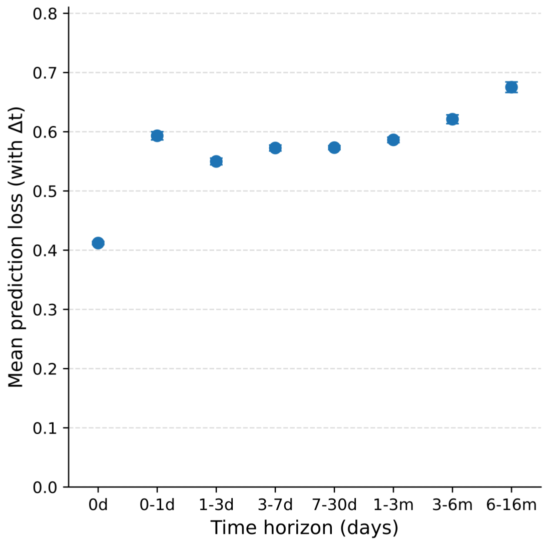

**Fig. S7 | Prediction error across prediction horizons.**

Mean test-set prediction error (MSE) as a function of the time interval ($\Delta t$) between consecutive clinical visits, computed using the true $\Delta t$ values. Results are shown for binned prediction horizons ranging from 0 days to 6-16 months. Error bars represent the standard error of the mean.

### References

Goldberger, A. L., L. A. Amaral, L. Glass, et al. 2000. “PhysioBank, PhysioToolkit, and PhysioNet: Components of a New Research Resource for Complex Physiologic Signals.” *Circulation* 101 (23): E215–20.

Johnson, Alistair E. W., Lucas Bulgarelli, Lu Shen, et al. 2023. “MIMIC-IV, a Freely Accessible Electronic Health Record Dataset.” *Scientific Data* 10 (1): 1.

Liaw, Richard, Eric Liang, Robert Nishihara, Philipp Moritz, Joseph E. Gonzalez, and Ion Stoica. 2018. “Tune: A Research Platform for Distributed Model Selection and Training.” In *arXiv [cs.LG]*. July 13. arXiv. https://doi.org/10.48550/arXiv.1807.05118.
